# Markers of blood-brain barrier disruption increase early and persistently in COVID-19 patients with neurological manifestations

**DOI:** 10.1101/2022.09.26.22280358

**Authors:** V. Bonetto, L. Pasetto, I. Lisi, M. Carbonara, R. Zangari, E. Ferrari, V. Punzi, S. Luotti, N. Bottino, B. Biagianti, C. Moglia, G. Fuda, R. Gualtierotti, F. Blasi, C. Canetta, N. Montano, M. Tettamanti, G. Camera, M. Grimoldi, G. Negro, N. Rifino, A. Calvo, P. Brambilla, F. Biroli, A. Bandera, A. Nobili, N. Stocchetti, M. Sessa, E.R. Zanier

## Abstract

**Background:** Coronavirus disease 2019 (COVID-19) leads to peripheral and central disorders, frequently with neurological implications. Blood-brain barrier disruption (BBBd) has been hypothesized as a mechanisms in the acute phase. We tested whether markers of BBBd, brain injury and inflammation could help identify a blood signature for disease severity and neurological complications.

**Methods:** Biomarkers of BBBd (MMP-9, GFAP), neuronal damage (NFL) and inflammation (PPIA, IL-10, TNFα) were measured by SIMOA, AlphaLISA and ELISA, in two COVID-19 patient cohorts with high disease severity (ICU Covid; n=79) and neurological complications (NeuroCovid; n=78), and in two control groups with no COVID-19 history: healthy subjects (n=20) and patients with amyotrophic lateral sclerosis (ALS; n=51).

**Results:** Biomarkers of BBBd and neuronal damage were high in COVID-19 patients, with levels similar to or higher than in ALS. NeuroCovid patients had lower levels of PPIA but higher levels of MMP-9 than ICU Covid patients. There was evidence of different temporal dynamics in ICU Covid compared to NeuroCovid patients with PPIA and IL-10 levels highest in ICU Covid patients in the acute phase. In contrast, MMP-9 was higher in the acute phase in NeuroCovid patients, with severity-dependency in the long term. We also found clear severity-dependency of NFL and GFAP.

**Conclusions:** The overall picture points to an increased risk of neurological complications in patients with high levels of biomarkers of BBBd. Our observations may provide hints for therapeutic approaches mitigating BBBd to reduce the neurological damage in the acute phase and potential dysfunction in the long term.

## Introduction

SARS-CoV-2 infection is associated with neurological symptoms and complications that range from headache, anosmia and dysgeusia, to severe complications such as cerebrovascular events, encephalopathy, Guillain-Barré syndrome, and dementia-like syndrome [1]. In addition, many COVID-19 patients develop a ‘post-COVID-19 syndrome’ defined as the persistence of a wide spectrum of symptoms beyond four weeks after infection [2]. In symptomatic COVID-19 patients, a community-based study with over half a million people in the UK estimated that about one in three experienced at least one persistent symptom for 12 weeks or more [3]. In a population-based study in Lombardy, the post-COVID-19 condition was associated with death, rehospitalization and use of health resources [4]. Long-term neuropsychological impairments such as executive, attentional and memory deficits, are reported even after mild infection [5]. While the exact causes of post-COVID-19 syndrome remain largely elusive, the prevalence of associated neurological symptoms with an increased risk of anxiety and depression at 16-month follow-up [6] suggests a brain origin [7,8].

There is neurochemical evidence of neuronal injury in patients with COVID-19 [9,10], with reports of a severity-dependent increase of neurofilament light chain (NFL) at 4-month follow-up, further supporting ongoing brain injury even weeks and months after acute infection [11]. Not surprisingly, the neurological complications are associated with worse functional outcome, particularly in older subjects and those with comorbidities [12].

Hypotheses of pathogenic processes implicated in acute and delayed brain injury following a SARS-CoV-2 infection include: i) viral invasion, ii) bioenergy failure, iii) autoimmunity, and iv) innate neuroimmune responses [13]. In all these processes the blood-brain barrier (BBB), which maintains the specialized microenvironment of the neural tissue by regulating the trafficking of substances between the blood and brain compartments, has a central role.

Brain endothelial cells are the primary unit in close association with pericytes and astrocytes [14]. Pericytes, which are key cells in maintaining and supporting vascular homeostasis and barrier function[15], are also the main source of matrix metalloproteinase 9 (MMP-9) [16,17]. Inflammatory stimuli very rapidly activate MMP-9 at the pericyte somata, leading to degradation of the underlying tight junction complexes. Thus, MMP-9 can act as a toxic culprit of BBB disruption after acute [18,19] and neurodegenerative diseases [20]. Peptidyl prolyl cis-trans isomerase A (PPIA), also known as cyclophilin A, acts as an activator of MMP-9 [21,22] through binding to its CD147 receptor, which in addition has been proposed as an alternative route for SARS-CoV-2 infection [23].

Mechanistically, it has been demonstrated that the severe COVID-19-related cytokine storm is induced by a “spike protein-CD147-PPIA signaling axis” [24]. *In vivo* experiments using a preclinical mouse model indicated that an anti-CD147 antibody inhibited the cytokine storm of SARS-CoV-2 [24].

Astrocytic end-feet containing glial fibrillary acidic protein (GFAP) are an essential component of the BBB. High blood GFAP is a marker of structural damage in the acute phase of brain injury and a severity-dependent increase has been detected in COVID-19 patients [11,25]. These data highlight PPIA, MMP-9, and GFAP as key disease biomarkers, so their measurement in association with NFL, an established marker of brain injury, may help identify a blood signature for disease severity and neurological complications in COVID-19 patients (NeuroCovid).

We identified significant effects associated with SARS-CoV-2 infection in COVID-19 patients, with NeuroCovid subjects showing the highest levels of biomarkers associated with BBB disruption, while patients in the intensive care unit (ICU Covid) had higher levels of inflammatory response biomarkers.

## Materials and Methods

### Study approval

The study was approved by the ethics committees of the clinical centers involved: Fondazione IRCCS Ca’ Granda Ospedale Maggiore Policlinico, Milano (approval #868_2020, 28.10.2020), ASST Papa Giovanni XXIII, Bergamo (approval #123/20, 14.05.2020). Written consent was obtained from patients themselves or their legal representatives when they lacked capacity to consent. Wherever possible, informed consent was collected verbally. However, in most cases, due to the patient’s inability to provide informed consent or to collect it in compliance with the contagion prevention measures, the principle of secondary use of data was used in accordance with art. 28, paragraph 2, letter b) of the November 20, 2017 law, no. 167, included in the legislative decree 196/03 of art. 110-bis.

### Study populations

Two COVID-19 populations, referred to as ICU Covid and NeuroCovid, were recruited between February 2020 and February 2021. All participants received a positive PCR test for SARS-CoV-2 RNA on nasopharyngeal swab. Control groups free from COVID-19 history were patients with amyotrophic lateral sclerosis (ALS) and a healthy population. Their main demographic and clinical characteristics are reported in Table 1.

**Table 1.** Demographic and clinical characteristics of the patient cohorts.

| **Characteristics** | **ICU Covid** | **NeuroCovid** | **ALS** | **Healthy** |
| --- | --- | --- | --- | --- |
| N | 79 | 78 | 51 | 20 |
| Age at sampling, years, median (IQR) | 65<br>(58-70) | 61<br>(53-71) | 67<br>(62-71) | 61<br>(58-63) |
| Sex (% males) | 77% | 73% | 51% | 35% |
| Hospitalization, days, median (IQR) | 17<br>(10-28) | 29<br>(10-51) | - | - |
| Mortality (% deceased) | 34% | 16% | - | - |
| PaO<sub>2</sub>/FIO<sub>2</sub> ratio, median (IQR) | 131<br>(93-180) | - | - | - |
| ALSFRS-R<sup>1</sup> at sampling, median (IQR) | - | - | 33<br>(22-38) | - |

<sup>1</sup>ALSFRS-R: Revised Amyotrophic Lateral Sclerosis Functional Rating Scale

#### ICU Covid

All patients admitted to the ICU, Rianimazione 1 Fiera Milano COVID-19 (Fondazione IRCCS Ca’ Granda Ospedale Maggiore Policlinico, Milan, Italy) were screened for eligibility. Inclusion criteria for this study population were: i) signed informed consent and ii) >18 years of age. Exclusion criteria were: i) known previous neurological conditions; ii) more than 48h in another ICU before admission; iii) pregnancy. Out of 296 screened patients, 79 were recruited for the study.

#### NeuroCovid

Patients admitted to the COVID-19 wards (ASST Papa Giovanni XXIII, Bergamo, Italy) with neurological manifestations confirmed by a neurological consultation/neurophysiological assessment/neuroradiologic investigation were recruited. Patients’ samples had been collected in an observational study on neurological manifestations in COVID-19 patients approved by the local Ethics Committee (257/2020, 13/5/2020) [26]. The main neurological diagnoses in this cohort are summarized in Supplementary Table 1 in the Supplementary material. Inclusion criteria were: i) signed informed consent; ii) > 18 years of age; iii) cognitive or neurological symptoms presenting during COVID-19 hospitalization, for which a neurological consultation/neurophysiological assessment/neuroradiologic investigation was required; iv) blood samples available. Of the 137 NeuroCovid patients, 78 fitted these criteria and their samples were included in the study. Patients were stratified on the basis of the clinical outcome: discharged fully recovered (moderate), discharged with sequalae (severe), and deceased (dead).

#### ALS patients and healthy controls

Informed written consent was obtained from all subjects involved and the study was approved by the ethics committee of Azienda Ospedaliero Universitaria Città della Salute e della Scienza, Turin. Healthy subjects and ALS patients had no COVID-19 history. The diagnosis of ALS was based on a detailed medical history and physical examination and confirmed by electrophysiological evaluation. Inclusion criteria for ALS patients were: i) >18 years old; ii) diagnosis of definite, probable or laboratory-supported probable ALS, according to revised El Escorial criteria. Exclusion criteria were: i) diabetes or severe inflammatory conditions; ii) active malignancy; iii) pregnancy or breast-feeding.

### Study design

The study design is summarized in Figure 1. Two COVID-19 populations and ALS and healthy control groups were included (see *Study Populations* above). In the ICU Covid cohort, blood samples were drawn acutely at ICU admission (T0) and after 7 (T7) and 14 (T14) days. Clinical data were collected throughout the ICU stay and CT scans were done every two weeks when feasible. For the NeuroCovid cohort, the blood samples had initially been collected for clinical and not experimental purposes, so the samples available did not precisely match those collected in the ICU Covid cohort; therefore, we retrieved available samples from week 1 to week 2 in the ward (acute: T0-T14) and from longer timepoints (long term: T15-T90). Clinical data were retrieved from medical records. Blood samples and clinical analyses were then done at the Istituto di Ricerche Farmacologiche Mario Negri IRCCS.

**Figure 1:**
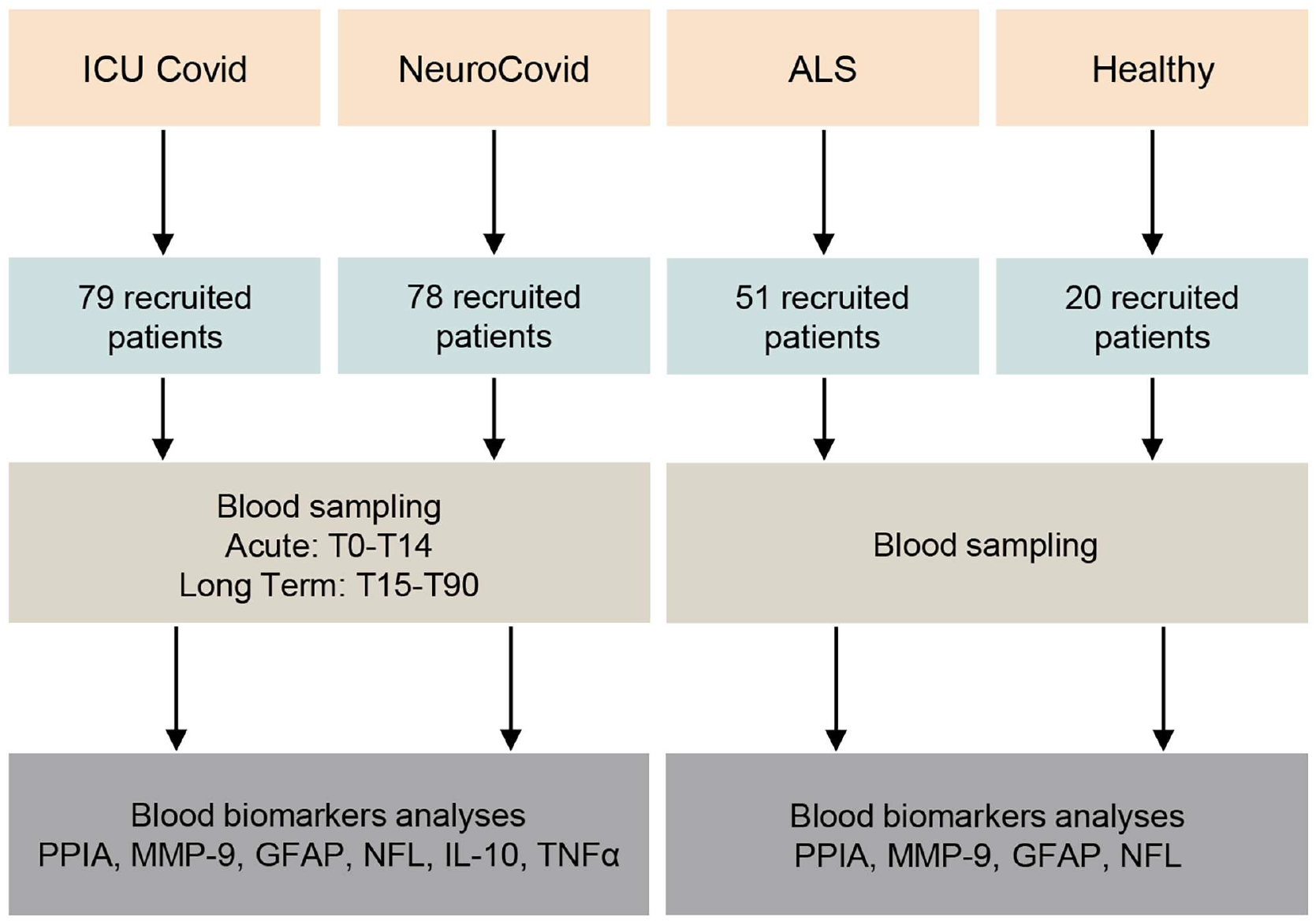
Schematic workflow for the biomarker characterization in two cohorts of COVID-19 patients and two cohorts of controls (ALS and healthy). The two COVID-19 cohorts were patients admitted to the ICU ward Rianimazione 1 Fiera Milano COVID-19 (ICU Covid; n=79) and to COVID-19 wards at the ASST Papa Giovanni XXIII Bergamo, with neurological complications (NeuroCovid; n=78). Blood samples were drawn acutely at ICU admission and after 7-14 days (T0-T14), and in the longer term between 15 and 90 days in the ward (T15-T90). Plasma samples were isolated, then analyzed for PPIA, MMP-9, GFAP, NFL, IL-10 and TNFα biomarkers. Control groups were ALS patients (n=51) and healthy subjects (n=20). Plasma samples were isolated, then analyzed for PPIA, MMP-9, GFAP and NFL.

### Biomarker analysis

Bloods were processed at the contributing centers and plasma samples were aliquoted, cryopreserved at -80°C and shipped to the Istituto di Ricerche Farmacologiche Mario Negri IRCCS for biomarker analyses. Levels of NFL, GFAP, IL-10 and TNFα were measured using commercially available single molecule array assay kits on an SR-X Analyzer (Neuro 2-Plex B (#103520), interleukin-10 (IL-10) (#101643) and tumor necrosis factor (TNFα) (#101580) advantage kits) as described by the manufacturer (Quanterix, Billerica, MA). A single batch of reagents was used for each analyte. MMP-9 was measured with an AlphaLISA kit for the human protein (#AL3138, PerkinElmer). AlphaLISA signals were measured using an Ensight Multimode Plate Reader (PerkinElmer). PPIA was measured with an ELISA for the human protein (#RD191329200R, BioVendor).

### Other laboratory data

Clinical and outcome data were retrieved from medical records for all patients.

### Statistical analysis

For each variable the differences between experimental groups were analysed by a Mann Whitney test or Kruskall-Wallis test, followed by Dunn’s post-hoc tests. Two-way ANOVA for repeated measures followed by Sidak’s post-hoc test was used to analyse biomarkers in ICU Covid patients. *P* values below 0.05 were considered significant. Prism 8.0 (GraphPad Software Inc., San Diego, CA) was used.

### Data availability

All data produced in the present study are available upon reasonable request to the authors.

## Results

### Blood biomarkers of BBB disruption and neuronal damage are high in COVID-19 patients with levels similar to or higher than in a severe neurodegenerative disease

PPIA, an inducer of MMP-9 [22] and cytokine storm [24], showed the highest levels in ICU Covid patients (**Fig. 2A**), while MMP-9, which is strictly related to BBB disruption, is highest in NeuroCovid patients (**Fig. 2B**). Both PPIA and MMP-9 in hospitalized COVID-19 patients are substantially higher than ALS patients and healthy controls (**Fig. 2A-B**). Plasma concentrations of GFAP were also high, irrespective of the neurological complications compared to healthy controls and were equal to or higher than the levels in ALS patients (**Fig. 2C**). NFL has similar behavior, with the highest levels in NeuroCovid significantly higher than in ICU Covid patients (**Fig. 2D**). These data suggest a clear neurological implication and call for a granular description of biomarker changes in these patient cohorts in relation to time and severity.

**Figure 2:**
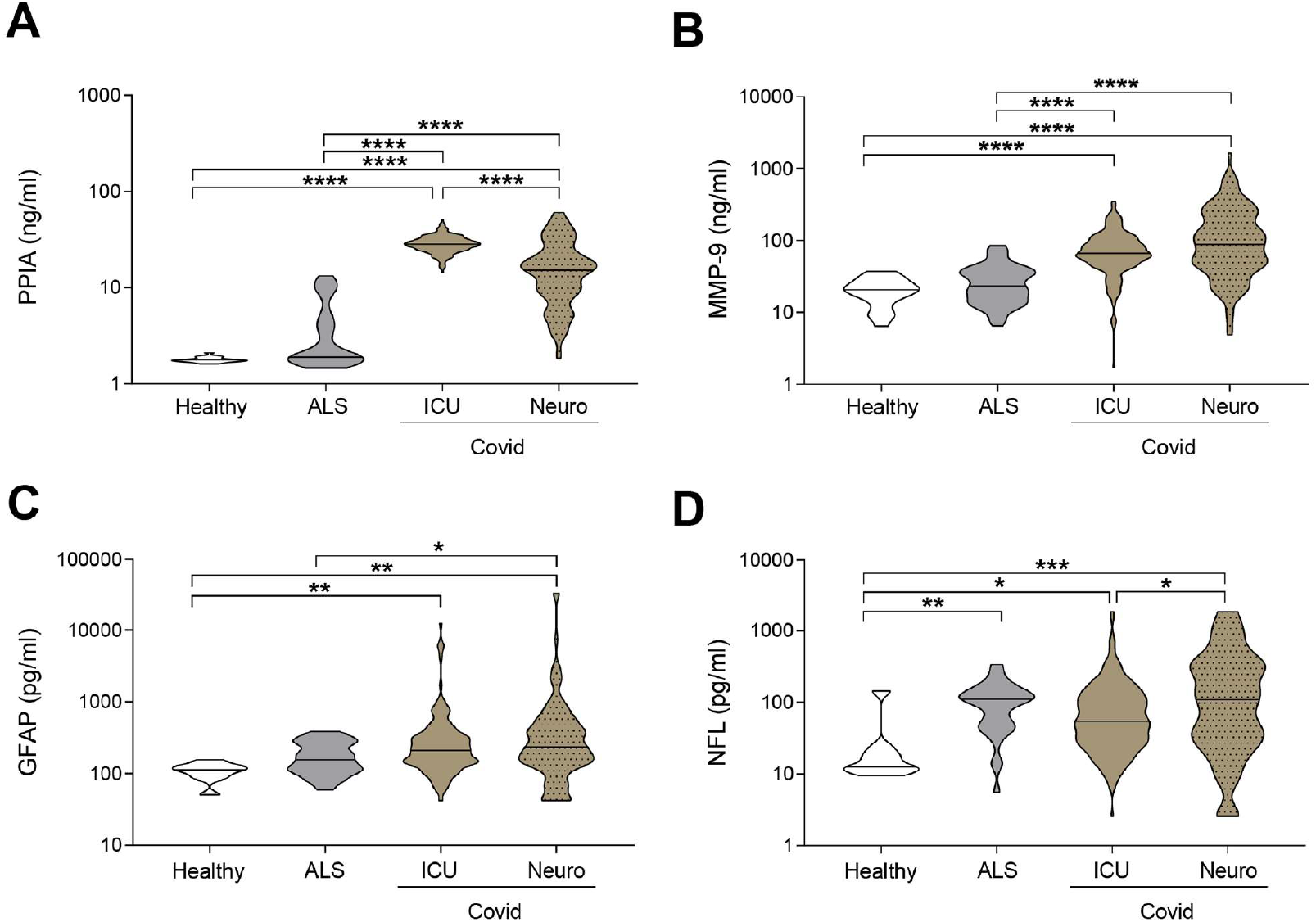
Comparison of biomarkers in COVID-19 and a neurodegenerative disorder. **(A-D)** PPIA **(A)**, MMP-9 **(B)**, GFAP (**C**), and NFL **(D)** concentrations were measured in plasma samples from ICU Covid patients (ICU n=196), NeuroCovid patients (n=120), ALS patients (PPIA n=50; MMP-9 n=51; GFAP and NFL n=34;) and healthy controls (PPIA n=18; MMP-9 n=20; GFAP and NFL n=9). Violin plots indicate median, variability and probability density of biomarker concentrations. **(A, B, D)** Kruskal-Wallis, p < 0.0001; (C) Kruskal-Wallis, p < 0.001. **(A-D)** *p < 0.05, **p < 0.01, ***p < 0.001, ****p < 0.0001 by Kruskal-Wallis, Dunn’s post hoc test.

### NeuroCovid patients have lower levels of the cytokine storm inducer PPIA but higher levels of BBB disruption markers

We characterized the severity-dependent changes and temporal dynamics of PPIA, MMP-9, GFAP and NFL in ICU Covid and NeuroCovid patients. In the acute phase, ICU Covid patients had higher PPIA levels than NeuroCovid patients (**Fig. 3A**). Among ICU Covid patients, a slight temporal increase was observed in the deceased group, leading to higher PPIA levels at 14 days than in alive patients (T14, **Fig. 3B**). NeuroCovid patients showed no severity dependency in the acute and the longer phases (**Fig. 3C, D**). In the acute phase, ICU Covid patients had lower MMP-9 levels than NeuroCovid patients (**Fig. 3E**). In the ICU Covid cohort, there was a slight decrease over the first two weeks in alive patients, leading to significantly lower levels on day 14 compared to deceased patients (T14, **Fig. 3F**). In the NeuroCovid cohort, MMP-9 levels were similarly high in alive and deceased patients in the acute phase, while in the longer term they showed severity dependency (**Fig. 3G-H**).

**Figure 3:**
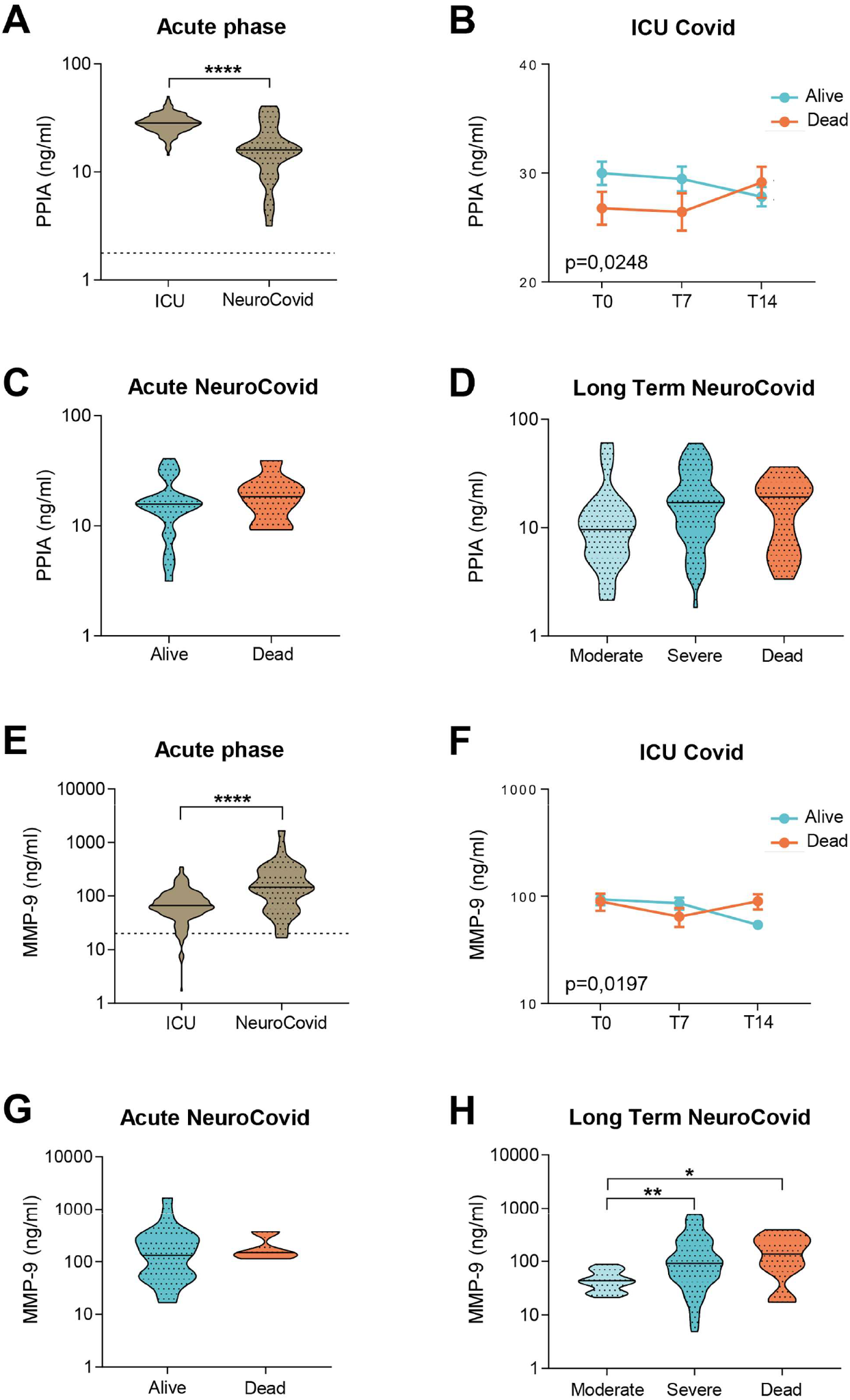
PPIA and MMP-9 in plasma of two cohorts of COVID-19 patients. **(A-H)** PPIA **(A-D)** and MMP-9 **(E-H)** concentrations were measured respectively by ELISA and AlphaLISA in plasma samples from two cohorts of COVID-19 patients. **(A, E)** Violin plots of PPIA **(A)** and MMP-9 **(E)** in the acute phase, in ICU Covid (n=196) and NeuroCovid samples (n=33). Dotted line indicates the mean levels in healthy controls. **(A, E)** Mann Whitney, ****p < 0.0001. **(B, F)** PPIA **(B)** and MMP-9 **(F)** were measured in ICU Covid patients, at ICU admission (T0) and after 7 (T7) and 14 days (T14). ICU Covid patients were stratified as alive (n=32) or dead (n=14). Data (mean ± SEM) indicate biomarker concentrations. **(B)** Two-way ANOVA for repeated measures, p < 0.05; **(F)** two-way ANOVA for repeated measures, p < 0.05. **(C, G)** PPIA **(C)** and MMP-9 **(G)** were measured in the acute phase in samples from NeuroCovid patients, stratified as alive (n=25) or dead (n=8). **(C)** Mann Whitney, p = 0.3966; **(G)** Mann Whitney, p = 0.5498. **(D, H)** The concentrations of PPIA **(D)** and MMP-9 **(H)** in the longer term, in samples from NeuroCovid patients, stratified as moderate (n=18), severe (n=58) or dead (n=11). **(D)** Kruskal-Wallis, p = 0.1175. **(H)** Kruskal-Wallis, p < 0.005; *p < 0.05, **p < 0.01 by Kruskal-Wallis, Dunn’s post hoc test.

In the acute phase, GFAP levels did not differ between groups (**Fig. 4A**). The longitudinal trajectories in ICU Covid patients showed an increase only in deceased patients, with the largest difference at admission (T0, **Fig. 4B**). NeuroCovid patients had widely varying GFAP levels in the acute phase (**Fig. 4A**). This is due to GFAP severity dependency, significant in the acute phase and as a tendency in the longer term **(Fig. 4C-D**).

**Figure 4:**
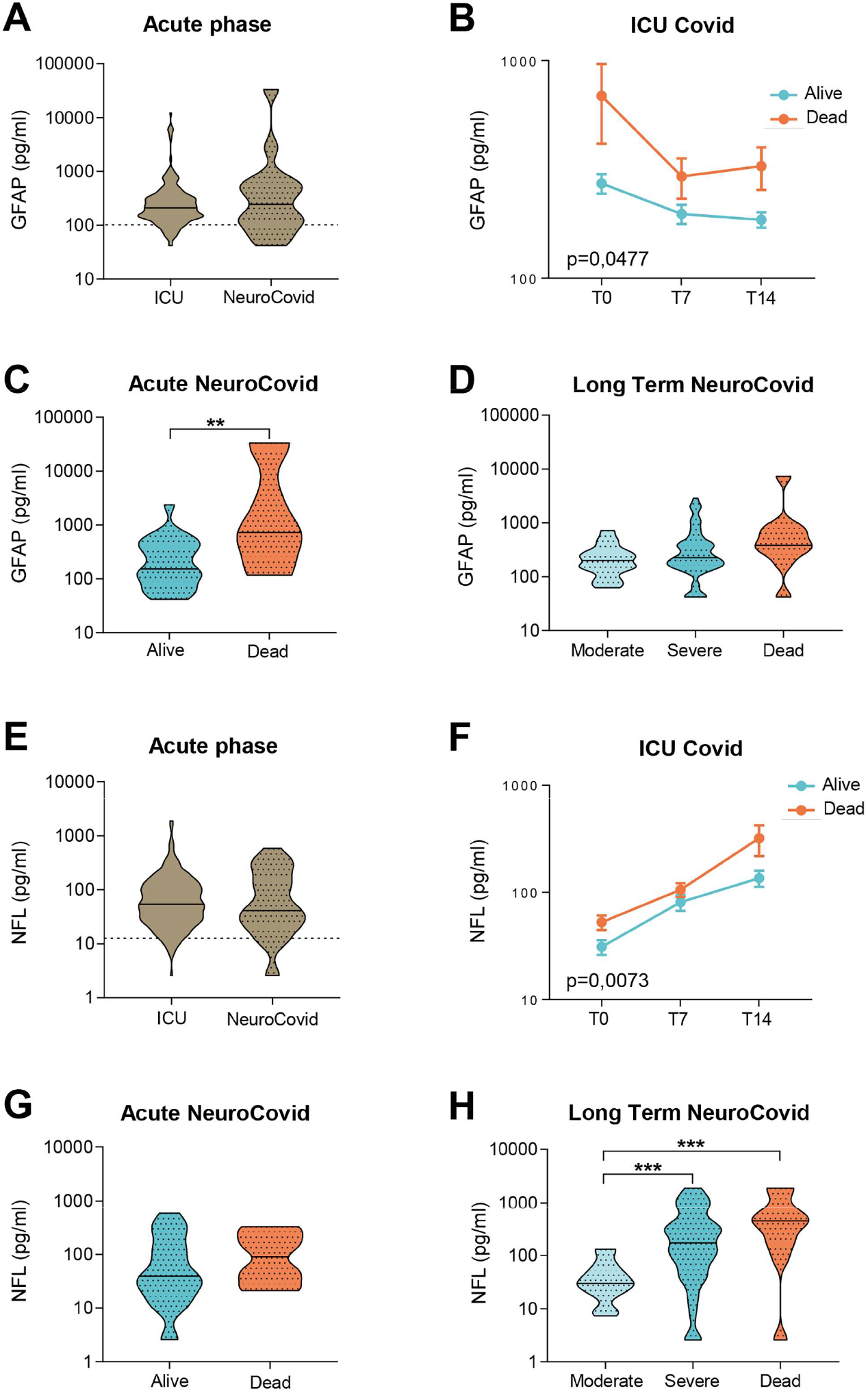
GFAP and NFL in plasma of two cohorts of COVID-19 patients. **(A-H)** GFAP **(A-D)** and NFL **(E-H)** concentrations were measured with Simoa technology in plasma from two cohorts of COVID-19 patients. **(A, E)** Violin plots of GFAP **(A)** and NFL **(E)** in the acute phase, in ICU Covid (n=196) and NeuroCovid samples (n=33). Dotted line indicates the mean level of healthy controls. **(A)** Mann Whitney, p = 0.7910; **(E)** Mann Whitney, p = 0.7054. **(B, F)** GFAP **(B)** and NFL **(F)** were measured in ICU Covid patients, at ICU admission (T0) and after 7 (T7) and 14 days (T14). ICU Covid patients were stratified as alive (n=32) or dead (n=14). Data (mean ± SEM) indicate biomarker concentrations. **(B)** Two-way ANOVA for repeated measures, p < 0.05; **p < 0.005 alive versus dead at T0 by Sidak’s post hoc test; **(F)** Two-way ANOVA for repeated measures, p < 0.01; ***p < 0.001 alive versus dead at T14 by Sidak’s post hoc test. **(C, G)** GFAP **(C)** and NFL **(G)** were measured in the acute phase in samples from NeuroCovid patients, stratified as alive (n=25) or dead (n=8). **(C)** Mann Whitney, **p < 0.01; **(G)** Mann Whitney, p = 0.2868. **(D, H)** The concentrations of GFAP **(D)** and NFL **(H)** in long-term samples from NeuroCovid patients, stratified as moderate (n=18), severe (n=58) or dead (n=11). **(D)** Kruskal-Wallis, p = 0.0570; **(H)** Kruskal-Wallis, p < 0.0001. ***p < 0.001 by Dunn’s post hoc test.

In the acute phase, ICU Covid patients had NFL levels similar to NeuroCovid patients (**Fig. 4E**). The trajectories of live and dead ICU Covid cohorts show a steep increase in NLF levels over the first two weeks, reaching the highest value for patients who were deceased 14 days from ICU admission (T14, **Fig. 4F**). While in the acute phase live and dead NeuroCovid patients had similar NFL levels **(Fig. 4G**), in the longer term NFL levels showed clear severity dependency (**Fig. 4H**).

Inflammatory markers of the systemic immune response, including IL-10 and TNFα, were also measured. In the acute phase, IL-10 levels were highest in ICU Covid compared to NeuroCovid patients (**Fig. 5A**), with a clear increase in deceased ICU Covid patients by day 14 (T14, **Fig. 5B**). In the NeuroCovid cohort IL-10 levels were similar in alive and deceased patients (**Fig. 5C**). In the longer term, however, severity dependency was observed (**Fig. 5D**).

**Figure 5:**
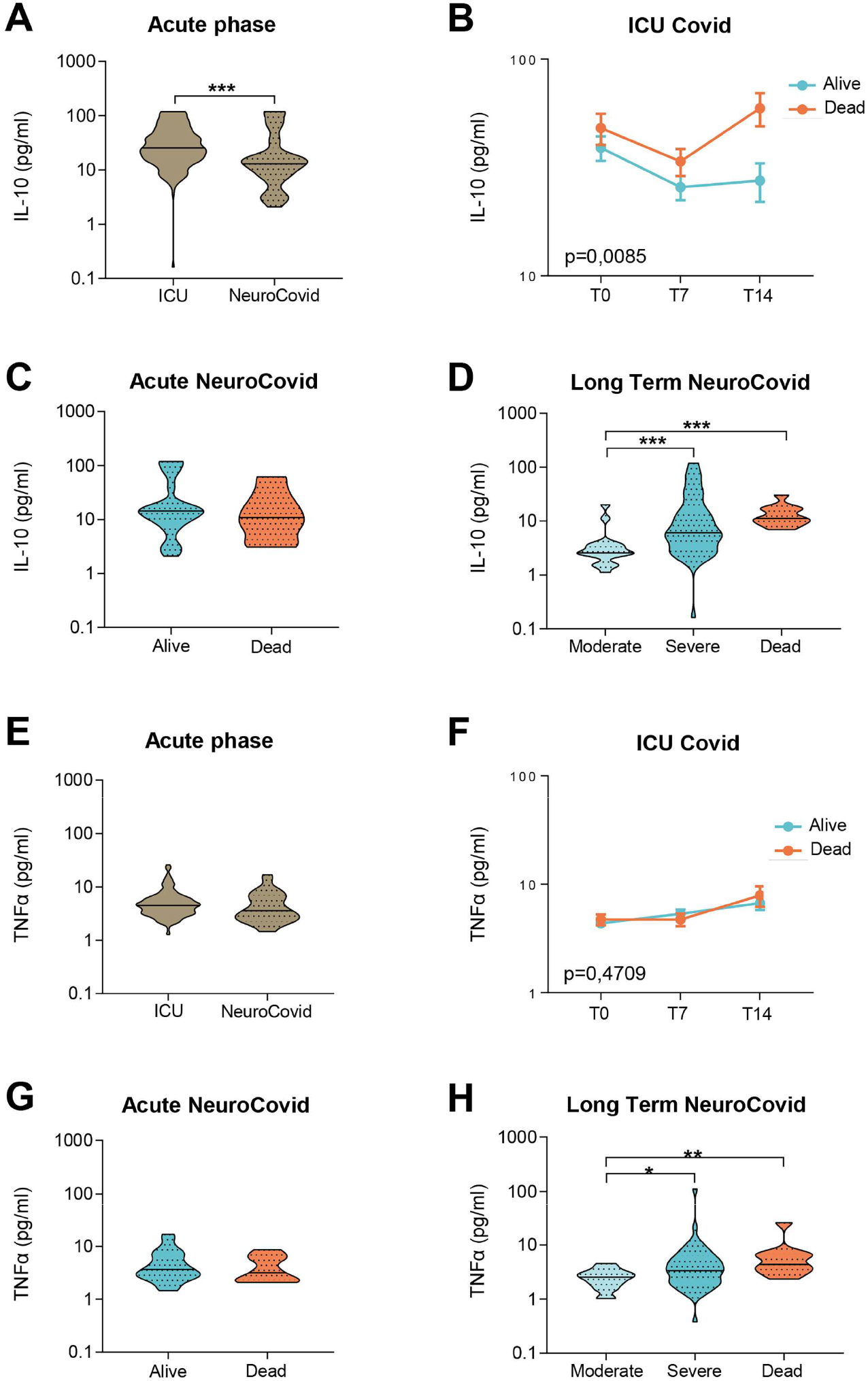
IL-10 and TNFα in plasma of two cohorts of COVID-19 patients. **(A-H)** IL-10 **(A-D)** and TNFα **(E-H)** concentrations were measured by Simoa technology in plasma from two cohorts of COVID-19 patients. **(A, E)** Violin plots of IL-10 **(A)** and TNFα **(E)** in the acute phase, in ICU Covid (n=196) and NeuroCovid samples (n=33). **(A)** Mann Whitney, ***p < 0.001. **(E)** Mann Whitney, p = 0.085. **(B, F)** IL-10 **(B)** and TNFα **(F)** were measured in ICU Covid patients at ICU admission (T0) and after 7 (T7) and 14 days (T14). ICU Covid patients were stratified as alive (n=32) or dead (n=14). Data (mean ± SEM) indicate biomarker concentrations. **(B)** Two-way ANOVA for repeated measures, p < 0.01 for cohort factor; **p < 0.005 alive versus dead at T14 by Sidak’s post hoc test. **(F)** Two-way ANOVA for repeated measures, p = 0.4709. **(C, G)** IL-10 **(C)** and TNFα **(G)** were measured in the acute phase in samples from NeuroCovid patients, stratified as alive (n=25) or dead (n=8). **(C)** Mann Whitney, p = 0.6652; **(G)** Mann Whitney, p = 0.5498. **(D, H)** The concentrations of IL-10 **(D)** and TNFα **(H)** at a longer time, in samples from NeuroCovid patients, stratified as moderate (n=18), severe (n=58) or dead (n=11). **(D)** Kruskal-Wallis, p < 0.0001; ***p < 0.001 by Kruskal-Wallis, Dunn’s post hoc test. **(H)** Kruskal-Wallis, p < 0.01; *p < 0.05, **p < 0.01 by Kruskal-Wallis, Dunn’s post hoc test.

In the acute phase, TNFα levels did not differ between ICU Covid and NeuroCovid patients (**Fig. 5E**) and within the ICU Covid cohort there were no temporal changes in alive and deceased patients up to day 14 (T14, **Fig. 5F**). In the NeuroCovid cohort, acute TNFα levels were similar in alive and deceased patients **(Fig. 5G**) but in the long term NeuroCovid patients showed significant severity dependency (**Fig. 5H**).

## Discussion

This study examined the effects of SARS-CoV-2 infection on blood biomarkers of BBB disruption, neuronal damage and systemic inflammation by longitudinally monitoring two patient cohorts of COVID-19, with increasing disease severity and neurological complications. Blood biomarkers of BBB disruption were elevated in COVID-19 patients with levels comparable to or even higher than in ALS patients, pointing to neurological implications over a range of disease severities.

There was evidence of different temporal dynamics in ICU Covid compared to NeuroCovid patients with PPIA, the potent activator of the cytokine storm and MMP-9 inducer, and IL-10, the master regulator of immunity to infection, with the highest levels in ICU Covid patients in the acute phase (**Fig. 6**). In contrast, MMP-9 was significantly higher in the acute phase in NeuroCovid patients, with severity dependency in the longer term. In line with previous findings, we found also clear severity dependency of NFL and GFAP levels with the highest levels in deceased patients, and severe NeuroCovid patients showing a tendency to maintain higher values than moderate patients in the longer term.

**Figure 6:**
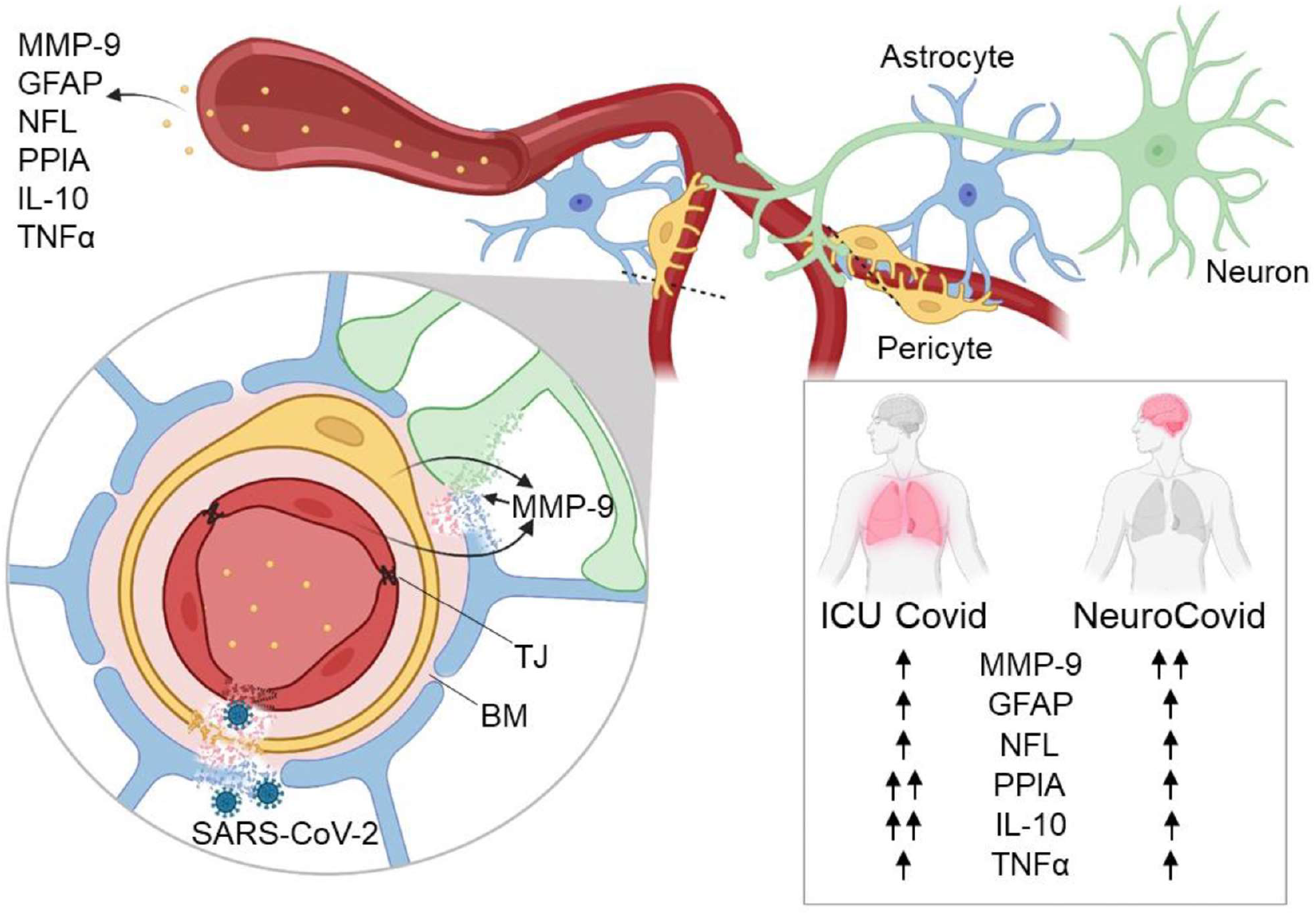
Highlights of the results. The effects of SARS-CoV-2 infection on blood biomarkers of BBB disruption (MMP-9, GFAP), neuronal damage (NFL) and systemic inflammation (PPIA, IL-10, TNFα) were measured in patients with severe disease (ICU Covid) and with neurological complications (NeuroCovid). Levels of PPIA and IL-10 in ICU Covid were higher than in NeuroCovid patients, while MMP-9 was significantly higher in NeuroCovid patients. Over-activation of MMP-9 may lead to degradation of tight junctions (TJ), basement membrane (BM) and laminin, implying BBB disruption, and penetration of SARS-CoV-2 into the brain and neuronal damage. Blood biomarkers of BBB disruption and neuronal damage were elevated in all COVID-19 patients, suggesting potential neurological dysfunctions in the long term, over a range of disease severities. Figure created with BioRender.com.

PPIA is a foldase and a molecular chaperone with multiple functions and substrates, including viral proteins essential for coronavirus replication [27]. Under certain conditions, such as inflammation, oxidative stress, and ischemia, PPIA is secreted extracellularly by several types of cells, including pericytes, vascular smooth muscle cells and macrophages, behaving as a pro-inflammatory cytokine, with potent chemotactic activity toward leukocytes [21]. Through the interaction with its CD147 receptor, in a NF-κB-dependent pathway, PPIA is an inducer of MMP-9 and of pro-inflammatory cytokines and chemokines [22,28]. High levels of PPIA have been seen in biofluids of several conditions associated with inflammation, including neurological and cardiovascular diseases [22,29]. Interestingly, high plasma concentrations of PPIA have also recently been reported in COVID-19 patients with mechanistic evidence for its involvement in the induction of the cytokine storm by activating CD147 [24].

A growing body of clinical data suggests that the cytokine storm is associated with COVID-19 severity, ICU admission, and is a crucial cause of death [30]. In agreement with this, our ICU Covid patients had PPIA concentrations substantially higher than NeuroCovid patients. Also noteworthy is the extremely high PPIA plasma concentration in all COVID-19 patients. This may be linked to its up-regulation upon interaction of SARS-CoV-2 with CD147, as observed in animal models [24], and may favor viral replication [27,31]. Similarly, MMP-9 was very high in all COVID-19 patients. However, NeuroCovid patients had the highest levels of MMP-9 in the acute phase, with persistent high levels in most severe patients in the long term. MMP-9 is a metalloproteinase with a wide substrate spectrum and is an important mechanism for fine-tuning cellular processes, but if aberrantly activated it is a key factor in BBB disruption and neuronal damage, by degrading tight junction proteins and laminin [18–20]. MMP-9 can be induced by inflammatory signaling cascades with CD147 acting as the major upstream inducer in the CNS [32]. CD147 is highly expressed in the brain capillary endothelium and various sub-regions of the brain [33]. Brain pericytes are the main source of MMP-9 at the neurovascular unit and it is rapidly released in response to inflammatory stimuli [16,17]. It has also been demonstrated *in vitro* and *in vivo* that SARS-CoV-2 can infect the brain microvascular endothelial cells and cross the BBB by MMP-9-mediated disruption of basement membrane [34]. Therefore, one can hypothesize that a local, early high MMP-9 concentration at the neurovascular unit in NeuroCovid patients, rather than extensive systemic inflammation as in ICU Covid patients, may be responsible for the BBB disruption that triggers neurological complications following SARS-CoV-2 infection.

Astrocytic end feet cover more than 99% of the neurovascular surface and directly affect BBB permeability [35]. GFAP is a highly expressed protein of the CNS, almost exclusively in astrocytes. In neuropathological conditions, GFAP is released into the bloodstream either by direct venous drainage or through a compromised BBB [36]. Blood GFAP can therefore serve as a useful biomarker and prognostic tool for numerous neurological conditions [25].

While classically considered a marker of astrogliosis, the presence of glial-derived proteins in peripheral body fluids has been suggested as indicating BBB disruption in acute CNS injury [37]. In the case of traumatic brain injury, it has been recently suggested that high blood GFAP concentrations might reflect damage to astrocytic end feet enveloping the BBB, thus releasing GFAP directly into the blood when the BBB is injured [38]. Elevated GFAP plasma levels have been reported in COVID-19 patients [10,38] and were in line with neuropathological data indicating post-mortem evidence of BBB disruption and gliosis [39]. In accordance with this, here we report high GFAP levels in a severity-dependent manner, with significantly higher levels at acute timepoints in deceased patients. GFAP only tended to be higher in NeuroCovid patients than in ICU Covid patients. However, the NeuroCovid cohort included several patients with Guillain-Barré syndrome in which blood-nerve-barrier (BNB) disruption is a key step [40]. BNB lacks astrocytes and glia limitans, so the detection of barrier damage through GFAP in these cases is underestimated.

NFL is an established marker of axonal injury [41]. Although axonal degeneration is not a specific feature of ALS, NFL is considered its most characteristic biomarker since its concentration is higher than in any other neurological disease [42,43]. This may be because neurofilaments are abundantly expressed in the large myelinated axons involved in the degenerative process, which is particularly fast and severe in ALS compared to other diseases. The only other condition in which the NFL plasma concentration is as high as in ALS is HIV-associated dementia (HAD) [44]. Interestingly, it seems that HIV-related CNS degeneration starts during primary infection and continues during subsequent stages of the disease. However, CSF NFL levels in primary infection are associated with CNS immune activation and BBB disruption but are not accompanied by high CSF total tau and low amyloid beta peptides, as in subjects with HAD [45]. This indicates that this early neuronal injury is less severe and/or involves a different mechanism and can in fact be halted by antiretroviral therapy [46]. Plasma NFL levels were high in all COVID-19 patients, with NeuroCovid patients reaching the same high levels as in HAD [44]. Although the overall picture points to an increased risk for neurological dysfunctions in the long term, the mechanism and extent to which acute axonal damage, in combination with systemic inflammation and BBB disruption, can predispose to neurodegeneration calls for further investigation.

Our observations may provide hints for a preventive approach. Should further evidence confirm that the neuronal damage found is secondary to, or exacerbated by, BBB disruption, therapies reducing BBB damage could serve as a valuable aid in attenuating the neurological damage in the acute phase and potential dysfunction in the longer term. Interestingly, MMP-9 stands as a druggable target since a set of potent MMPs inhibitors are already available for clinical use [47], furthermore drugs targeting the PPIA-CD147-MMP-9 signaling pathway are also under investigation. A PPIA inhibitor, cyclosporine A (CsA), a well-known immunosuppressive drug, and Meplazumab, an anti-CD147 monoclonal antibody, are being assessed in clinical trials up to phase 2/3 (NCT05113784) for the treatment of severe COVID-19 [48]. There are some indications from observational studies of milder COVID-19 and lower mortality in solid organ transplant recipients and autoimmune disease patients under CsA treatment [49].

Patients in our study were recruited before the vaccination campaign. Luckily, vaccination with at least two doses of COVID-19 vaccine has been shown to substantially reduce the most common post-acute COVID-19 symptoms [50]. Since COVID-19 vaccination may also have a protective effect against the post-COVID-19 syndrome follow-up studies are now needed to monitor distinct long-term consequences in vaccinated and non-vaccinated subjects.

## Data Availability

All data produced in the present study are available upon reasonable request to the authors

## Abbreviations

BBB: blood-brain barrier
MMP-9: matrix metalloproteinase 9
PPIA: peptidyl prolyl cis-trans isomerase A
GFAP: glial fibrillary acidic protein
NFL: neurofilament light chain
ALS: amyotrophic lateral sclerosis
IL-10: interleukin-10
TNFα: tumor necrosis factor α

## Acknowledgments

We would like to thank the “COVID-19 NETWORK” WORKING GROUP. Fondazione IRCCS Ca’ Granda Ospedale Maggiore Policlinico. <u>Scientific Direction</u>: Silvano Bosari, Luigia Scudeller, Giuliana Fusetti, Laura Rusconi, Silvia Dell’Orto; <u>Department of Transfusion Medicine and Hematology (Biobank)</u>: Daniele Prati, Luca Valenti, Silvia Giovannelli, Maria Manunta, Giuseppe Lamorte, Francesca Ferarri; <u>Infectious Diseases Unit</u>: Andrea Gori, Alessandra Bandera, Antonio Muscatello, Davide Mangioni, Laura Alagna, Giorgio Bozzi, Andrea Lombardi, Riccardo Ungaro, Giuseppe Ancona, Gianluca Zuglian, Matteo Bolis, Nathalie Iannotti, Serena Ludovisi, Agnese Comelli, Giulia Renisi, Simona Biscarini, Valeria Castelli, Emanuele Palomba, Marco Fava, Valeria Fortina, Carlo Alberto Peri, Paola Saltini, Giulia Viero, Teresa Itri, Valentina Ferroni,Valeria Pastore,Roberta Massafra,Arianna Liparoti,Toussaint Muheberimana, Alessandro Giommi, Rosaria Bianco, Rafaela Montalvao De Azevedo, Grazia Eliana Chitani; <u>Angelo Bianchi Bonomi Hemophilia and Thrombosis Center and Fondazione Luigi Villa</u>: Flora Peyvandi, Roberta Gualtierotti, Barbara Ferrari, Raffaella Rossio, Nadia Boasi, Erica Pagliaro, Costanza Massimo, Michele De Caro, Andrea Giachi; <u>UOC Internal Medicine, Immunology and Allergology:</u> Nicola Montano, Barbara Vigone, Chiara Bellocchi, Angelica Carandina, Elisa Fiorelli, Valerie Melli, Eleonora Tobaldini; <u>Respiratory Unit and Cystic Fibrosis Adult Center:</u> Francesco Blasi, Stefano Aliberti, Maura Spotti,Leonardo Terranova, Sofia Misuraca, Alice D’Adda, Silvia Della Fiore, Marta Di Pasquale, Marco Mantero Martina Contarini, Margherita Ori, Letizia Morlacchi, Valeria Rossetti, Andrea Gramegna, Maria Pappalettera, Mirta Cavallini, Agata Buscemi; <u>Cardiology Unit:</u> Marco Vicenzi, Irena Rota. <u>Emergency Unit</u>: Giorgio Costantino, Monica Solbiati, Ludovico Furlan, Marta Mancarella, Giulia Colombo, Giorgio Colombo, Alice Fanin, Mariele Passarella; <u>Acute Internal Medicine</u>: Valter Monzani, Ciro Canetta, Angelo Rovellini, Laura Barbetta, Filippo Billi, Christian Folli, Silvia Accordino; <u>Rare Diseases Center</u>: Diletta Maira, Cinzia Maria Hu, Irene Motta, Natalia Scaramellini; <u>General Medicine and Metabolic Diseases:</u> Anna Ludovica Fracanzani, Rosa Lombardi, Annalisa Cespiati ; <u>Geriatric Unit</u>: Matteo Cesari,Tiziano Lucchi,Marco Proietti, Laura Calcaterra, Clara Mandelli, Carlotta Coppola, Arturo Cerizza. Intensive Care Unit: Antonio Maria Pesenti, Giacomo Grasselli, Alessandro Galazzi. Istituto di Ricerche Farmacologiche Mario Negri IRCCS: Alessandro Nobili, Mauro Tettamanti, Igor Monti, Alessia Antonella Galbussera.

## Supplementary Material

**Supplementary Table 1.** Case definition for NeuroCovid cohort

|  |  |
| --- | --- |
| **NeuroCovid total patients (N)** | 78 |
| **Neurological complications** | N (%) |
| *Cerebrovascular disorders<sup>1</sup>* | 18 (23) |
| Ischemic stroke | 14 |
| Hemorrhagic stroke | 4 |
| Transient ischemic attack | 1 |
| *Peripheral neuropathies* | 26 (33) |
| Guillain Barré Syndrome<sup>2</sup> | 26 |
| *Encephalopathies/Encephalites<sup>3</sup>* | 26 (33) |
| *Miscellaneous* | 8 (10) |
| Epilepsy | 4 |
| Myelopathy | 1 |
| Syncope | 1 |
| Movement disorder | 1 |
| Headache | 1 |

<sup>1</sup>As defined by Sacco et al. (Sacco et al., 2013)
<sup>2</sup>As defined by Sejvar et al. (Sejvar et al., 2011)
<sup>3</sup>As defined by Quist-Paulsen et al. (Quist-Paulsen et al., 2019)

**Supplementary Table 2.**

| Characteristics |  | PPIA<br>(ng/ml) | MMP-9<br>(ng/ml) | GFAP<br>(pg/ml) | NFL<br>(pg/ml) | IL-10<br>(pg/ml) | TNFα<br>(pg/ml) |
| --- | --- | --- | --- | --- | --- | --- | --- |
| **ICU Covid** | N | 196 | 196 | 196 | 196 | - | - |
|  | Median | 28.3 | 66.3 | 210.2 | 54.5 | - | - |
|  | IQR | 24.8-31.9 | 45.7-99.1 | 140.9-341.3 | 27.0-119.9 | - | - |
|  | Figure | 2A | 2B | 2C | 2D | - | - |
| **NeuroCovid** | N | 120 | 120 | 120 | 120 | - | - |
|  | Median | 15.2 | 88.4 | 233.0 | 109.2 | - | - |
|  | IQR | 8.4-22.4 | 44.4-206.9 | 140.0-472.8 | 29.8-335.7 | - | - |
|  | Figure | 2A | 2B | 2C | 2D | - | - |
| **ALS** | N | 50 | 51 | 34 | 34 | - | - |
|  | Median | 1.9 | 23.3 | 155.4 | 111.3 | - | - |
|  | IQR | 1.8-4.9 | 14.6-37.3 | 109.1-268.7 | 45.6-136.8 | - | - |
|  | Figure | 2A | 2B | 2C | 2D | - | - |
| **Healthy** | N | 18 | 20 | 9 | 9 | - | - |
|  | Median | 1.8 | 20.6 | 112.8 | 12.6 | - | - |
|  | IQR | 1.7-1.9 | 13.9-27.0 | 89.2-129.6 | 11.4-21.5 | - | - |
|  | Figure | 2A | 2B | 2C | 2D | - | - |
| **Acute ICU Covid** | N | 196 | 196 | 196 | 196 | 196 | 196 |
|  | Median | 28.3 | 66.3 | 210.2 | 54.5 | 25.4 | 4.5 |
|  | IQR | 24.8-31.9 | 45.7-99.1 | 140.9-341.3 | 27.0-119.9 | 15.5-47.5 | 3.2-6.0 |
|  | Figure | 3A | 3E | 4A | 4E | 5A | 5E |
| **Acute NeuroCovid** | N | 33 | 33 | 33 | 33 | 33 | 33 |
|  | Median | 16.0 | 146.4 | 244.1 | 41.1 | 13.0 | 3.6 |
|  | IQR | 10.8-21.4 | 61.2-279.5 | 105.5-583.8 | 21.6-155.6 | 7.6-24.1 | 2.4-6.6 |
|  | Figure | 3A | 3E | 4A | 4E | 5A | 5E |
| **Acute ICU Covid alive** | N | 32 | 32 | 32 | 32 | 32 | 32 |
|  | IQR | 27.8-30.0 | 54.3-93.8 | 185.8-272.6 | 31.0-135.8 | 25.6-39.0 | 4.4-6.7 |
|  | Figure | 3B | 3F | 4B | 4F | 5B | 5F |
| **Acute ICU Covid dead** | N | 14 | 14 | 14 | 14 | 14 | 14 |
|  | Median | 26.8 | 89.4 | 327.4 | 105.9 | 48.1 | 4.7 |
|  | IQR | 26.4-29.2 | 64.7-90.0 | 293.6-690.0 | 52.7-320.1 | 33.7-59.4 | 4.7-7.9 |
|  | Figure | 3B | 3F | 4B | 4F | 5B | 5F |
| **Acute NeuroCovid alive** | N | 25 | 25 | 25 | 25 | 25 | 25 |
|  | Median | 15.8 | 131.8 | 152.8 | 39.1 | 14.5 | 3.7 |
|  | IQR | 10.1-17.8 | 50.2-279.5 | 82.6-410.3 | 17.2-142.4 | 8.3-28.3 | 2.6-6.7 |
|  | Figure | 3C | 3G | 4C | 4G | 5C | 5G |
| **Acute NeuroCovid dead** | N | 8 | 8 | 8 | 8 | 8 | 8 |
|  | Median | 18.4 | 148.0 | 723.1 | 89.7 | 10.9 | 3.2 |
|  | IQR | 11.1-23.5 | 120.9-307.3 | 245.8-25835 | 28.6-254.1 | 4.5-27.3 | 2.2-6.3 |
|  | Figure | 3C | 3G | 4C | 4G | 5C | 5G |
| **Long Term NeuroCovid moderate** | N | 18 | 18 | 18 | 18 | 18 | 18 |
|  | Median | 9.6 | 43.5 | 199.0 | 29.8 | 2.6 | 2.6 |
|  | IQR | 5.4-15.0 | 26.2-65.3 | 131.6-290.6 | 19.1-55.2 | 2.1-3.6 | 1.7-3.1 |
|  | Figure | 3D | 3H | 4D | 4H | 5D | 5H |
| **Long Term NeuroCovid severe** | N | 58 | 58 | 58 | 58 | 58 | 58 |
|  | Median | 17.1 | 91.4 | 227.2 | 172.4 | 6.1 | 3.4 |
|  | IQR | 9.2-29.8 | 43.7-209.3 | 154.2-396.0 | 57.9-363.2 | 3.3-16.3 | 2.1-6.0 |
|  | Figure | 3D | 3H | 4D | 4H | 5D | 5H |
| **Long Term NeuroCovid dead** | N | 11 | 11 | 11 | 11 | 11 | 11 |
|  | Median | 19.2 | 136.7 | 382.1 | 449.0 | 11.2 | 4.4 |
|  | IQR | 5.4-25.6 | 68.9-245.8 | 277.2-849.1 | 161.3-770.6 | 9.1-17.1 | 3.3-6.6 |
|  | Figure | 3D | 3H | 4D | 4H | 5D | 5H |

